# Low-pass whole genome sequencing is a reliable and cost-effective approach for copy number variant analysis in the clinical setting

**DOI:** 10.1101/2023.05.26.23290606

**Authors:** Patricia C. Mazzonetto, Darine Villela, Silvia Souza da Costa, Ana C. V. Krepischi, Fernanda Milanezi, Michele P. Migliavacca, Paulo M. Pierry, Adriano Bonaldi, Luiz Gustavo D. Almeida, Camila Alves De Souza, José Eduardo Kroll, Marcelo G. Paula, Rodrigo Guarischi-Sousa, Cristovam Scapulatempo-Neto, Carla Rosenberg

## Abstract

We evaluated the performance of low-pass whole genome sequencing (LP-WGS) to detect copy number variants (CNVs) in clinical cytogenetics. DNA samples with known CNVs detected by chromosomal microarray analyses (CMA) were selected for comparison; our panel included 44 DNA samples (12 prenatal and 32 postnatal), comprising a total of 55 chromosome imbalances. The selected cases were chosen to provide a wide range of clinically relevant CNVs, being the vast majority associated with intellectual disability or recognizable syndromes. The chromosome imbalances ranged in size from 75 kb to 90.3 Mb, including aneuploidies and two cases of mosaicism. All CNVs were successfully detected by LP-WGS, showing a high level of consistency and robust performance of the sequencing method. Notably, the size of chromosome imbalances detected by CMA and LP-WGS were compatible between the two different platforms, which indicates that the resolution and sensitivity of the LP-WGS approach are at least similar to those provided by CMA. Our data show the potential use of LP-WGS to detect CNVs in clinical diagnosis and confirm the method as an alternative for chromosome imbalances detection. The diagnostic effectiveness and feasibility of LP-WGS, in this technical validation study, were evidenced by a clinically representative dataset of CNVs that allowed a systematic assessment of the detection power and the accuracy of the sequencing approach. Further, since the software used in this study is commercially available, the method can be easily tested and implemented in a routine diagnostic setting.

## INTRODUCTION

Copy number variants (CNVs) are a common source of genetic variation that has been implicated in many genomic disorders, Mendelian diseases, and complex traits ^1–3^. Chromosomal microarray analysis (CMA), including array-comparative genomic hybridization (array-CGH) and SNP-array, are the gold standard procedure to detect CNVs in the clinical setting ^4, 5^. Nonetheless, next generation sequencing (NGS) is an alternative state-of-art technology, allowing detection of genetic alterations with unprecedent level of resolution. In fact, a higher sensitivity and resolution for CNV detection is achieved through paired-end whole genome sequencing using deep coverage (>30x), although its cost is still considerably higher than microarrays ^6^. With the dropping in sequencing costs in the last years, an approach that is rapidly being adopted as an alternative method for CNV analysis in several medical centres is the low-pass whole genome sequencing (LP-WGS) (coverage 0.1-1x) ^7^. Not surprisingly, LP-WGS is cheaper, faster, technically easier to implement and automate in molecular diagnostics in comparison to CMA. By reducing the sequencing coverage while increasing its resolution, as is the case of LP-WGS, it is possible to detect chromosomal abnormalities with high precision. The current most important applications of LP-WGS includes non-invasive prenatal testing (NIPT) ^8–10^, pre-implantation genetic testing (PGT) ^11, 12^, liquid biopsy ^13, 14^, and solid tumour analysis ^15, 16^.

Emerging evidence have supported the performance of LP-WGS for detecting CNVs in clinical cytogenetics, particularly, in prenatal and postnatal diagnosis ^17–23^. Dong et al. demonstrated that chromosomal disorders or microdeletion/microduplication syndromes can be effectively detected using a high resolution genome sequencing method ^19^. The authors pointed out the potential use of LP-WGS to facilitate genetic diagnosis in prenatal and postnatal samples that were not detected by karyotyping and/or CMA. This evidence was reinforced by other investigations, which showed that CNV analysis from LP-WGS in some cases outperformed the CMA method depending on the microarray platform being used in the clinical setting ^20^. However, the sequencing depth (0.1-1x) and mode (single-end or paired-end) varies between studies, making data replication difficult. Moreover, data analysis consist of in-house pipelines that are hard to implement in a diagnostic routine and requires bioinformatics skills. In this study, we aimed to investigate the efficiency of LP-WGS to detect CNVs in prenatal and postnatal samples using sequencing services and a commercial software; the protocol was designed so that it could be easily tested and implemented in a clinical laboratory routine.

## MATERIALS AND METHODS

### Sample selection

The DNA samples selected for this study were obtained from 44 unrelated individuals previously referred to molecular investigation in clinical cytogenetics. The patients were primarily investigated by CMA (either array-CGH or SNP-array), currently considered the gold standard diagnostic test for CNV analysis. Our DNA panel included 12 prenatal and 32 postnatal samples, comprising a total of 55 genomic imbalances. All these DNA alterations used as positive controls in the LP-WGS experiments are described in **Table 1**. The CNVs were chosen mainly to represent a wide range of clinically relevant CNVs detected by CMA in our diagnostic routine, being the vast majority of them associated with intellectual disability or recognizable syndromes. The selected CNVs (1) contained at least one coding sequence, (2) were mapped to a variety of chromosomes, (3) ranged in copy number state from zero to 3/4, and (4) ranged in genomic size from 75 kb to 90.3 Mb, including aneuploidies and two mosaic cases. Particularly, for methodology evaluation and quality control metrics, we used DNA extracted from different types of biological samples. Prenatal samples included chorionic villi, amniotic fluid, and miscarriage tissues, whereas postnatal samples included peripheral blood, blood from FTA card, and oral swab.

**Table 1.** Panel of CNVs previously detected in prenatal and postnatal samples by CMA used for the LP-WGS study.

| Sample | Sex | Type of sample | Cytoband | Alteration | Size (bp) | CNV Classification | Platform | Regions and known syndromes |
| --- | --- | --- | --- | --- | --- | --- | --- | --- |
| *A. Prenatal Samples* |  |  |  |  |  |  |  |  |
| 1 | F | Amniotic fluid | 2p11.2 | duplication | 215,493 | VUS | CytoSNP 850K | - |
| 2 | F | Amniotic fluid | 11q14.3 | duplication | 1,013,989 | VUS | CytoSNP 850K | - |
| 3 | F | Amniotic fluid | 22q11.21 | deletion | 2,210,039 | P | CytoSNP 850K | DiGeorge syndrome |
| 4 | M | Amniotic fluid | 12p13.33p11.1 | mosaic duplication | 34,522,878 | P | CytoSNP 850K | Pallister-Killian syndrome |
| 5 | F | Amniotic fluid | 21p11.2q22.3 | trisomy | 37,346,423 | P | CytoSNP 850K | Down syndrome/Trisomy 21 |
| 6 | F | Chorionic villus | 15q26.3 | terminal duplication | 1,059,572 | LP | CytoSNP 850K | - |
| 7 | F | Chorionic villus | 21p11.2q22.3 | trisomy | 37,308,667 | P | CytoSNP 850K | Down syndrome/Trisomy 21 |
| 8 | F | Chorionic villus | 18p11.32q23 | trisomy | 80,169,792 | P | CytoSNP 850K | Edwards syndrome/Trisomy 18 |
| 9 | M | Miscarriage | 21p11.2q22.3 | trisomy | 37,308,667 | P | CytoSNP 850K | Down syndrome/Trisomy 21 |
| 10 | M | Miscarriage | 22p11.2q13.33 | trisomy | 40,003,915 | P | CytoSNP 850K | Trisomy 22 |
| 11 | F | Miscarriage | 16p13.3q24.3 | trisomy | 90,170,122 | P | CytoSNP 850K | Trisomy 16 |
| 12 | F | Miscarriage | 16p13.3q24.3 | trisomy | 90,170,122 | P | CytoSNP 850K | Trisomy 16 |
| *B. Postnatal Samples* |  |  |  |  |  |  |  |  |
| 13 | M | Blood | 17p13.3p13.1 | mosaic deletion | 4,421,240 | P | Agilent 180k | - |
| 14 | M | Blood | 15q11.2 | deletion | 53,715 | VUS | Agilent 180k | - |
| 15 | F | Blood | 8p12q12.1 | duplication | 26,658,507 | P | Agilent 60k | - |
| 16 | M | Blood | Xq24 | deletion | 131,840 | P | Agilent 60k | - |
| 17 | F | Blood | 22q11.21 | deletion | 2,178,919 | P | Agilent 60k | DiGeorge syndrome |
| 18 | F | Blood | 4p16.3 | terminal deletion | 3,767,489 | P | Agilent 60k | Wolf-Hirschhorn syndrome |
|  |  |  | 7p22.3p22.1 | terminal duplication | 67,827,615 | P |  | - |
| 19 | M | Blood | 3p26.3p25.3 | terminal deletion | 10,513,502 | P | CytoSNP 850K | Chromosome 3pter-p25 deletion syndrome |
|  |  |  | 3q28q29 | terminal duplication | 8,954,943 | P |  | Chromosome 3q29 duplication syndrome |
| 20 | F | Blood | 7q11.23 | deletion | 1,483,526 | P | Agilent 180k | Williams-Beuren syndrome |
| 21 | M | Blood | 3p25.1 | duplication | 159,827 | LB | Agilent 180k | - |
| 22 | M | Blood | 15q13.3 | duplication | 493,316 | LB | CytoSNP 850K | 15q13.3 recurrent region (includes *CHRNA7* and *OTUD7A*) |
| 23 | F | Blood | 16p13.11 | duplication | 1,323,380 | P | Agilent 180k | 16p13.11 recurrent region (BP2-BP3) (includes *MYH11*) |
| 24 | M | Blood | 1q21.1 | deletion | 380,746 | VUS | Agilent 60k | 1q21.1 recurrent (TAR syndrome) region (proximal, BP2-BP3) (includes *RBM8A*) |
| 25 | M | Blood | 1p36.23p36.22 | deletion | 2,021,048 | P | Agilent 60k | 1p36 deletion syndrome |
| 26 | M | Blood | 17p11.2 | deletion | 3,033,705 | P | Agilent 60k | Smith-Magenis syndrome |
| 27 | M | Blood | 17p11.2 | duplication | 3,511,672 | P | Agilent 60k | Potocki-Lupsk syndrome (17p11.2 duplication syndrome) |
| 28 | M | Blood | 15q11.2q13.1 | deletion | 5,171,568 | P | Agilent 60k | Prader Willi/Angelman syndrome |
| 29 | M | Blood | 7q11.23 | duplication | 1,258,420 | P | Agilent 60k | Williams-Beuren duplication region syndrome / 7q11.23 duplication syndrome |
| 30 | F | Blood | 16p11.2 | deletion | 600,645 | P | CytoSNP 850K | 16p11.2 recurrent region (proximal, BP4-BP5) (includes *TBX6*) / Chromosome 16p11.2 deletion syndrome |
| 31 | M | Blood | Xq28 | duplication | 644,393 | P | Agilent 60k | Intellectual developmental disorder, X-linked syndromic, Lubs type/Xq28 region (includes *MECP2*) |
| 32 | F | Blood | 15q11.2 | deletion | 608,459 | P | Agilent 60k | Chromosome 15q11.2 deletion syndrome / 15q11.2 recurrent region (BP1-BP2) (includes NIPA1) |
| 33 | M | Blood | 3q21.2q21.3 | deletion | 4,222,858 | P | Agilent 180K | - |
|  |  |  | Xp22.31 | duplication | 1,612,520 | VUS |  | Xp22.31 recurrent region (include *STS* gene) |
| 34 | F | Blood | 5p15.33p15.1 | terminal deletion | 15,394,134 | P | CytoSNP 850K | Cri du chat syndrome |
|  |  |  | 5q34q35.3 | terminal duplication | 15,119,618 | P |  | includes 5q35 recurrent (Sotos syndrome) region (includes NSD1) |
|  |  |  | 10q24.2 | deletion | 243,133 | LB |  | - |
| 35 | F | Blood | 7p22.3p22.1 | terminal duplication | 6,378,080 | P | CytoSNP 850K | - |
|  |  |  | 22q13.2q13.33 | terminal deletion | 8,245,687 | P |  | - |
| 36 | F | Blood | 4p16.3p11 | terminal duplication | 48,665,733 | P | CytoSNP 850K | Includes 4p16.3 terminal region (Wolf-Hirschhorn syndrome) |
|  |  |  | 4p16.3 | terminal deletion | 99,170 | B |  | - |
|  |  |  | 10q21.3 | duplication | 274,685 | VUS |  | - |
| 37 | M | Blood | 9p24.3p21.1 | terminal duplication | 29,322,080 | P | CytoSNP 850K | - |
|  |  | Blood | 7q36.2q36.3 | terminal deletion | 5,991,645 | P | CytoSNP 850K | - |
| 38 | F | Blood | 15q11.2q13.1 | deletion | 4,876,948 | P | CytoSNP 850K | Prader-Willi/Angelman syndrome |
| 39 | F | Blood | 10q11.22q11.23 | duplication | 4,160,321 | P | CytoSNP 850K | - |
| 40 | M | Blood | 4p16.3 | terminal duplication | 3,988,435 | P | CytoSNP 850K | 4p16.3 terminal (Wolf-Hirschhorn syndrome) region |
|  |  |  | 4p23.3p23.1 | terminal deletion | 6,934,909 | P |  | - |
| 41 | F | Blood | Xq22.1q28 | terminal deletion | 55,600,111 | P | CytoSNP 850K | Includes Xq28 recurrent region (int22h1/int22h2-flanked) (includes *RAB39B*) and Xq25 region (includes STAG2) |
| 42 | F | Blood from FTA card | 10q21.1 | duplication | 131,931 | VUS | CytoSNP 850K | - |
| 43 | M | Oral swab | 15q11.1q13.1 | duplication/triplication | 8,432,793 | P | CytoSNP 850K | Chromosome 15q11-q13 duplication syndrome |
| 44 | F | Oral swab | 17p13.3 | duplication | 545,052 | P | CytoSNP 850K | 17p13.3 (Miller-Dieker syndrome) region (includes *YWHAE* and *PAFAH1B1*) |
|  |  |  | 17p13.3 | duplication | 391,655 | P |  |  |

CMA: chromosomal microarray analysis; LP-WGS: low-pass whole genome sequencing, F: female; M: male; VUS: variants of unknown significance, P: Pathogenic; LP: Likely Pathogenic; LB: Likely Benign; B: Benign; CNV: copy number variation.

### Chromosomal microarrays analysis (CMA)

SNP-array experiments were performed using the CytoSNP 850K BeadChip from Illumina (California, USA), and the array-CGH experiments were carried out using either 60K or 180K whole-genome platforms from Agilent Technologies (California, USA), following the manufacturers recommendations. Data were analyzed using the BlueFuse Multi v4.5 software (Illumina, USA) and Genomic Workbench 7.0 software (Agilent Technologies, USA) for SNP-array and array-CGH, respectively. A significance threshold of 5.0E-6 was applied for CNV calls. Log_2_ ratio and B Allele Frequency (BAF) (in the case of SNP-arrays) values were plotted along genomic coordinates, and the chromosome regions with either copy number or allele frequency alterations, were identified. SNP-array and array-CGH analysis were conducted according to parameters previously reported^24–26^.

### Low-pass whole genome sequencing (LP-WGS)

LP-WGS experiments were performed either on BGI (Pequim, China) or Illumina (California, USA) next generation sequencing (NGS) platforms. Briefly, the sequencing results generated ∼16 million paired-end reads of 150 bp per sample, corresponding to a 1x coverage. These reads were aligned to the human genome (GRCh38) using the BWA v0.7.17^27^ to generate the BAM files. Module MarkDuplicates from GATK^28^ was used to identify PCR duplicates and mark reads for exclusion in the downstream analysis. CNV data analysis was performed using the NxClinical software (BioDiscovery, California, USA), which calls copy number changes by comparing the number of reads of an experimental sample to an internal reference library, constructed based on whole genome data from controls samples sequenced at the same depth coverage. The data were normalized and the log_2_ ratio Test/Reference was calculated. Following the same principle of microarray analysis, the theoretically expected log2 ratio value, when there are no changes in copy number, corresponds to zero (test/reference = 1). To identify CNVs, we used the SNP-FASST2 segmentation algorithm, based on the Hidden Markov Model, with a sensitivity threshold of 1.0 E-6. A genomic segment was considered duplicated or deleted when the log_2_ ratio of a given region encompassing at least three targets was above 0.3 or below −0.3, respectively; further, we considered a mosaic duplication or deletion when the log_2_ ratio were above 0.1 or below −0.1, respectively, and encompassed at least three consecutive targets, as previously described^25, 26^.

### CNV clinical interpretation

Detected CNVs were classified according to the *European guidelines for constitutional cytogenomics analysis* ^4^, *American College of Medical Genetics* (ACMG) and *Clinical Genome Resource* (ClinGen) guidelines ^29^.

## RESULTS

To evaluate the performance of LP-WGS to detect CNVs in clinical cytogenetics, we used DNA samples with known CMA results for comparison. The genomic coordinates of the chromosome imbalances previously identified by CMA were compared to those derived from LP-WGS data (**Table 2)**. All CNVs detected by CMA were detected by LP-WGS. The comparisons of the CNV data extracted from CMA and LP-WGS showed that the calculated size of the genomic imbalances detected by these two methods were very similar, varying mostly according to the position of the probes in the microarray platform (**Table 2**). Genomic regions near centromeres and telomeres as well as some segmental duplications regions showed unspecific CNV calls in the LP-WGS data, as expected; these CNV calls were excluded from our analysis. Importantly, using the same criteria to call CNVs using microarray and sequencing data, no additional changes (false positive results) were observed in LP-WGS data. Overall, by using CMA results as reference, LP-WGS provided 100% sensitivity and 100% specificity for detecting CNVs, indicating a high level of consistency and robust performance of the NGS platform.

**Table 2.** Comparison of the CNV data extracted from CMA and LP-WGS

| Sample | Type of sample | Cytoband | Alteration | CMA (GRch38) |  |  | LP-WGS (GRch38) |  |  |
| --- | --- | --- | --- | --- | --- | --- | --- | --- | --- |
|  |  |  |  | Start | End | Size (bp) | Start | End | Size (bp) |
| A. Prenatal Samples |  |  |  |  |  |  |  |  |  |
| 1 | Amniotic fluid | 2p11.2 | duplication | 85,940,555 | 86,156,048 | 215,493 | 85,913,006 | 86,163,005 | 249,999 |
| 2 | Amniotic fluid | 11q14.3 | duplication | 89,969,500 | 90,983,489 | 1,013,989 | 89,730,391 | 90,980,390 | 1,249,999 |
| 3 | Amniotic fluid | 22q11.21 | deletion | 18,899,402 | 21,109,441 | 2,210,039 | 18,439,130 | 21,539,129 | 3,099,999 |
| 4 | Amniotic fluid | 12p13.33p11.1 | mosaic duplication | 82,453 | 34,605,331 | 34,522,878 | 1 | 35,268,550 | 35,268,549 |
| 5 | Amniotic fluid | 21p11.2q22.3 | trisomy | 9,363,560 | 46,709,983 | 37,346,423 | 8,156,379 | 46,709,983 | 38,553,604 |
| 6 | Chorionic villus | 15q26.3 | terminal duplication | 100,861,387 | 101,920,959 | 1,059,572 | 100,839,865 | 101,991,189 | 1,151,324 |
| 7 | Chorionic villus | 21p11.2q22.3 | trisomy | 9,371,576 | 46,680,243 | 37,308,667 | 8,157,439 | 46,709,983 | 38,552,544 |
| 8 | Chorionic villus | 18p11.32q23 | trisomy | 87,505 | 80,257,297 | 80,169,792 | 1 | 80,373,285 | 80,373,284 |
| 9 | Miscarriage | 21p11.2q22.3 | trisomy | 9,371,576 | 46,680,243 | 37,308,667 | 8,158,045 | 46,709,983 | 38,551,938 |
| 10 | Miscarriage | 22p11.2q13.33 | trisomy | 10,753,385 | 50,757,300 | 40,003,915 | 10,510,001 | 50,818,468 | 40,308,467 |
| 11 | Miscarriage | 16p13.3q24.3 | trisomy | 38,165 | 90,208,287 | 90,170,122 | 1 | 90,338,345 | 90,338,344 |
| 12 | Miscarriage | 16p13.3q24.3 | trisomy | 38,165 | 90,208,287 | 90,170,122 | 6 | 90,338,345 | 90338339 |
| B. Postnatal Samples |  |  |  |  |  |  |  |  |  |
| 13 | Blood | 17p13.3p13.1 | mosaic deletion | 2,952,205 | 7,373,445 | 4,421,240 | 2,951,517 | 7,401,516 | 4,449,999 |
| 14 | Blood | 15q11.2 | deletion | 24,869,158 | 24,922,873 | 53,715 | 24,864,883 | 24,939,829 | 74,946 |
| 15 | Blood | 8p12q12.1 | duplication | 32,283,366 | 58,941,873 | 26,658,507 | 31,960,001 | 58,752,722 | 26,792,721 |
| 16 | Blood | Xq24 | deletion | 119,459,408 | 119,591,248 | 131,840 | 119,486,412 | 119,750,407 | 263,995 |
| 17 | Blood | 22q11.21 | deletion | 18,907,307 | 21,086,226 | 2,178,919 | 18,689,130 | 21,389,129 | 2,699,999 |
| 18 | Blood | 4p16.3 | terminal deletion | 49,338 | 3,816,827 | 3,767,489 | 1 | 4,161,302 | 4,161,301 |
|  |  | 7p22.3p22.1 | terminal duplication | 149,248 | 67,976,863 | 67,827,615 | 1 | 6,862,396 | 6,862,395 |
| 19 | Blood | 3p26.3p25.3 | terminal deletion | 11,458 | 10,524,960 | 10,513,502 | 1 | 10,510,000 | 10,509,999 |
|  |  | 3q28q29 | terminal duplication | 191,886,101 | 200,841,044 | 8,954,943 | 189,085,425 | 198,295,559 | 9,210,134 |
| 20 | Blood | 7q11.23 | deletion | 73,238,378 | 74,721,904 | 1,483,526 | 73,215,839 | 74,865,838 | 1,649,999 |
| 21 | Blood | 3p25.1 | duplication | 12,591,863 | 12,751,690 | 159,827 | 12,590,527 | 12,765,962 | 175,435 |
| 22 | Blood | 15q13.3 | duplication | 31,727,716 | 32,221,032 | 493,316 | 31,689,865 | 32,439,864 | 749,999 |
| 23 | Blood | 16p13.11 | duplication | 14,874,998 | 16,198,378 | 1,323,380 | 14,810,001 | 16,510,000 | 1,699,999 |
| 24 | Blood | 1q21.1 | deletion | 145,635,445 | 146,016,191 | 380,746 | 145,372,530 | 146,334,587 | 962,057 |
| 25 | Blood | 1p36.23-p36.22 | deletion | 8,289,900 | 10,310,948 | 2,021,048 | 8,303,510 | 10,453,509 | 2,149,999 |
| 26 | Blood | 17p11.2 | deletion | 16,929,582 | 19,963,287 | 3,033,705 | 16,951,517 | 19,951,516 | 2,999,999 |
| 27 | Blood | 17p11.2 | duplication | 16,819,758 | 20,331,430 | 3,511,672 | 16,751,517 | 20,501,516 | 3,749,999 |
| 28 | Blood | 15q11.2-q13.1 | deletion | 21,242,091 | 26,413,659 | 5,171,568 | 23,389,862 | 28,339,864 | 4,950,002 |
| 29 | Blood | 7q11.23 | duplication | 73,308,984 | 74,567,404 | 1,258,420 | 73,315,839 | 74,715,838 | 1,399,999 |
| 30 | Blood | 16p11.2 | deletion | 29,603,655 | 30,204,300 | 600,645 | 29,560,001 | 30,160,000 | 599,999 |
| 31 | Blood | Xq28 | duplication | 153,467,182 | 154,111,575 | 644,393 | 153,876,737 | 154,326,736 | 449,999 |
| 32 | Blood | 15q11.2 | deletion | 22,612,562 | 23,221,021 | 608,459 | 22,264,861 | 23,014,861 | 750,000 |
| 33 | Blood | 3q21.2-q21.3 | deletion | 126,377,431 | 130,600,289 | 4,222,858 | 124,885,425 | 129,135,424 | 4,249,999 |
|  |  | Xp22.31 | duplication | 6,534,632 | 8,147,152 | 1,612,520 | 6,533,095 | 8,133,094 | 1,599,999 |
| 34 | Blood | 5p15.33p15.1 | terminal deletion | 38,141 | 15,432,275 | 15,394,134 | 1 | 15,460,000 | 15,459,999 |
|  |  | 5q34q35.3 | terminal duplication | 166,002,593 | 181,122,211 | 15,119,618 | 166,012,882 | 181,312,881 | 15,299,999 |
|  |  | 10q24.2 | deletion | 98,927,092 | 99,170,225 | 243,133 | 98,920,376 | 99,170,375 | 249,999 |
| 35 | Blood | 7p22.3p22.1 | terminal duplication | 44,935 | 6,423,015 | 6,378,080 | 1 | 6,237,395 | 6,237,394 |
|  |  | 22q13.2q13.33 | terminal deletion | 42,511,613 | 50,757,300 | 8,245,687 | 42,639,130 | 50,818,468 | 8,179,338 |
| 36 | Blood | 4p16.3p11 | terminal duplication | 149,953 | 48,815,686 | 48,665,733 | 135,000 | 49,611,924 | 49,476,924 |
|  |  | 4p16.3 | terminal deletion | 49,556 | 148,726 | 99,170 | 1 | 134,999 | 134,998 |
|  |  | 10q21.3 | duplication | 65,758,288 | 66,032,973 | 274,685 | 65,770,376 | 66,020,375 | 249,999 |
| 37 | Blood | 9p24.3p21.1 | terminal duplication | 46,587 | 29,368,667 | 29,322,080 | 1 | 29,310,000 | 29,309,999 |
|  | Blood | 7q36.2q36.3 | terminal deletion | 153,341,975 | 159,333,620 | 5,991,645 | 154,515,839 | 159,345,973 | 4,830,134 |
| 38 | Blood | 15q11.2q13.1 | deletion | 23,422,265 | 28,299,213 | 4,876,948 | 23,339,862 | 28,439,864 | 5,100,002 |
| 39 | Blood | 10q11.22q11.23 | duplication | 45,972,625 | 50,132,946 | 4,160,321 | 45,816,266 | 50,120,368 | 4,304,102 |
| 40 | Blood | 4p16.3 | terminal duplication | 49,556 | 4,037,991 | 3,988,435 | 1 | 4,020,606 | 4,020,605 |
|  |  | 4p23.3p23.1 | terminal deletion | 2,14,984 | 7,149,893 | 6,934,909 | 1 | 7,188,120 | 7,188,119 |
| 41 | Blood | Xq22.1q28 | terminal deletion | 100,406,971 | 156,007,082 | 55,600,111 | 100,429,595 | 155,676,736 | 55,247,141 |
| 42 | Blood from FTA card | 10q21.1 | duplication | 54,247,584 | 54,379,515 | 131,931 | 54,170,447 | 54,370,338 | 199,891 |
| 43 | Oral swab | 15q11.1q13.1 | duplication/triplication | 19,866,420 | 28,299,213 | 8,432,793 | 18,350,436 | 28,589,864 | 10,239,428 |
| 44 | Oral swab | 17p13.3 | duplication | 1,248,298 | 1,793,350 | 545,052 | 1,251,517 | 1,801,516 | 549,999 |
|  |  | 17p13.3 | duplication | 2,545,841 | 2,937,496 | 391,655 | 2,551,517 | 2,951,516 | 399,999 |

CMA: chromosomal microarray analysis; LP-WGS: low-pass whole genome sequencing.

**Figure 1** presents examples of CNVs detected by LP-WGS, using the NxClinical software. All cases with aneuploidies included in this validation study were detected on prenatal samples; a trisomy 16 found on a fetal tissue from miscarriage is shown in **Figure 1A**. In postnatal samples, all duplications and deletions ranging in different sizes were successfully identified. **Figure 1B** shows terminal deletion and duplication of the short and long arms of chromosome 5, respectively, which are evidenced by the log_2_ ratio profile. The 5.1 Mb deletion at 15q11.2q13.1 was observed in a patient with Angelman syndrome (**Figure 1C**); and the 1.4 Mb duplication at 7q11.23 in addition to the 10.2 Mb duplication at 15q11.1q13.1 (this alteration is evidenced by the log_2_ ratio as an amplification, suggesting the presence of four copies), correspond to clinically recognizable syndromes (**Figure 1D** and **1E**). The 4.4 Mb deletion at 17p13.3p13.1, although detected as a deletion, presents a log_2_ ratio not as low as other deletions (**Figure 1F**), suggesting mosaicism; indeed, this alteration was previously shown by FISH to be present in only approximately 50% of cultured lymphocytes^30^. To illustrate some of the small CNVs detected by LP-WGS, a 250 kb duplication at 10q21.3 and a deletion of the same size at 10q24.2 are shown on **Figure 1G** and **1H**, respectively.

**Figure 1.**
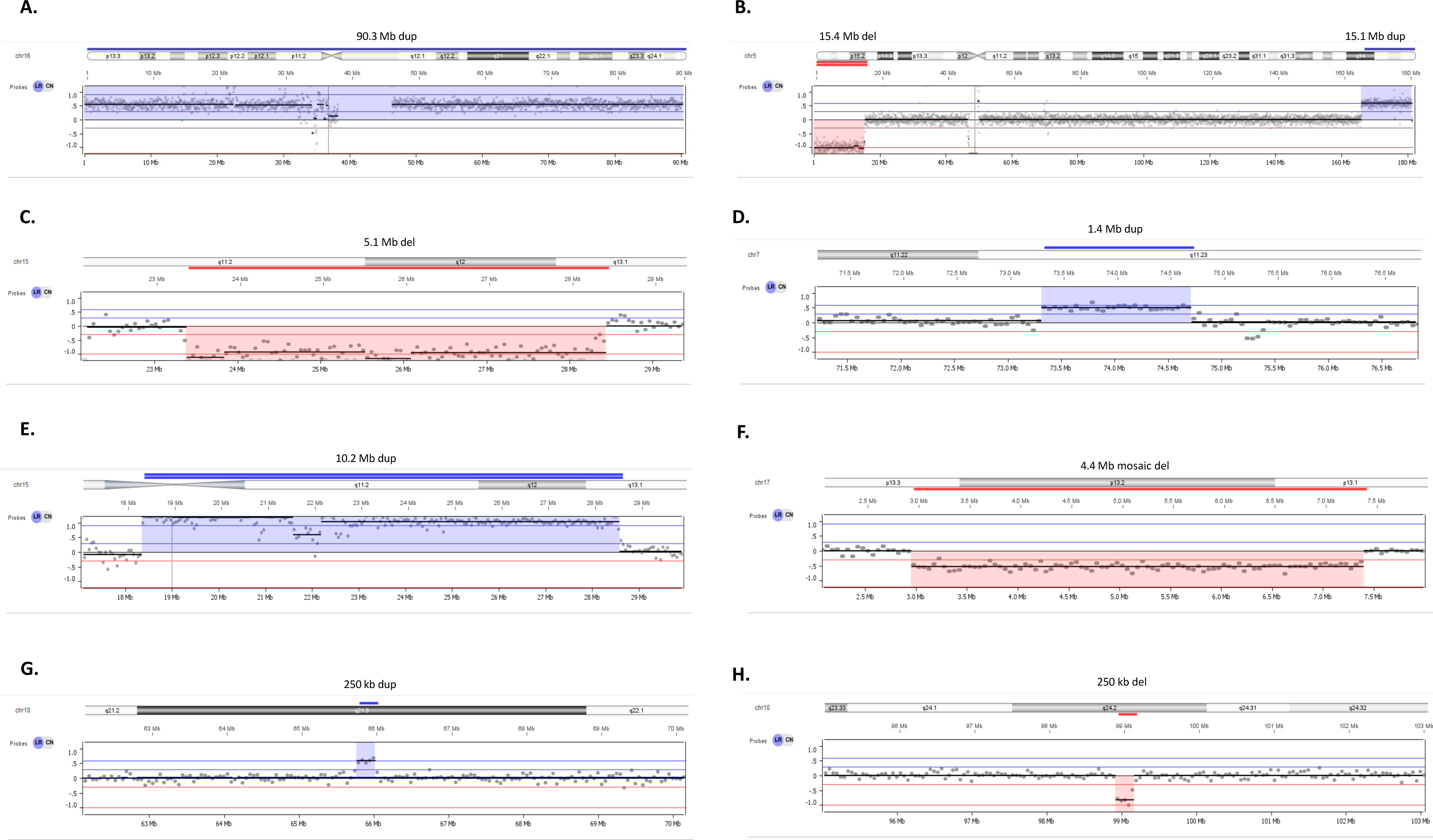
Copy number variants (CNVs) detected by low-pass whole genome sequencing (LP-WGS). The panel show eight different cases with previously detected CNVs here depicted using LP-WGS data, and its corresponding log_2_ ratio profile. (A) Log_2_ ratio profile of chromosome 16, showing a trisomy 16 (Sample 11); (B) Log_2_ ratio profile of chromosome 5, showing terminal duplication and deletion at the short and long arms of the chromosome, respectively (Sample 34); (C) Log_2_ ratio profile of the 15q11.2q13.1 cytobands, showing a 5.1 Mb deletion overlapping the Prader-Willi/Angelman syndrome region (Sample 38); (D) Log_2_ ratio profile of the 7q11.23 cytoband, showing a 1.5 Mb duplication (Sample 20), which correspond to the Williams-Beuren duplication region syndrome; (E) Log_2_ ratio profile of the 15q11.1q13.1 cytoband, showing a 10.2 Mb amplification, suggesting the presence of four copies (Sample 43) - this alteration is the clinically recognized syndrome 15q11-q13 duplication syndrome; (F) Log_2_ ratio profile of part of the short arm of chromosome 17 (Sample 13) showing an approximately 50% mosaic 4 Mb deletion at 17p13.3p13.1; (G) Log_2_ ratio profile of the 10q21.3 band showing a 250 kb duplication (Sample 35); and (H) Log_2_ ratio profile of the 10q24.2 cytoband showing a 250 kb deletion (Sample 34). Images were extracted from NxClinical software (Biodiscovery, USA). dup: duplication; del: deletion.

## DISCUSSION

Recent studies demonstrate that a high-resolution genome-wide sequencing approach can be an alternative method for CNV detection in routine clinical application^7, 17, 18, 20, 21, 31–33^. In this study, we aimed to evaluate the efficiency of LP-WGS to detect CNVs using a commercial software, in order to facilitate its implementation in diagnostic setting. The use of a clinically representative dataset of CNVs allowed a systematic assessment of detection power and accuracy of the sequencing approach; all numerical and structural chromosome alterations were successfully identified by LP-WGS, revealing consistency with CMA results. No difference was observed on the detection rate or accuracy between prenatal and postnatal samples.

The comparison of CNV data derived from microarrays and LP-WGS showed that the estimated sizes of chromosome imbalances detected by these two methods were very similar, indicating that the resolution and sensitivity of our approach is at least comparable to that of genomic microarrays. Small differences in size between array and sequencing data were due to the mapping of probes using different platforms. In fact, it has been showed that in some cases LP- WGS outperformed CMA in clinical cytogenetics ^20, 31^. Dong et al. developed an algorithm to map the precise CNV boundaries (windows) using an increment rate of coverage of the aligned reads, i.e., nonoverlapping windows ^31^. For any particular adjustable nonoverlapping window, the increment ratio of coverage was calculated as the coverage difference in that region. With this new established algorithm, the authors demonstrate that LP-WGS data provides a more uniform genome coverage, and it is more precise to identify critical regions of diseases when compared to CMA, which is limited by probe mapping and density. Particularly, the same research group showed that in the context of prenatal diagnosis, LP-WGS allowed detection of additional clinically relevant CNVs that were missed by CMA ^20^. The sequencing approach not only showed the advantage of identifying pathogenic CNVs present in regions with insufficient probe coverage on the microarray platform, but also demonstrated its increased sensitivity in detecting low-level of mosaicism (∼10%) ranging from large CNVs (>1.4 Mb) to partial aneuploidies (trisomy cases). Nonetheless, although the microarray platform used on those studies was considered to be reliable for prenatal diagnosis, its resolution is lower (60K) compared to other CMA platforms commercially available; hence, any comparison or extrapolation should be made with caution. In our prenatal samples, we applied the 850K SNP-array platform from Illumina, and it was not detected any additional finding in LP-WGS data (no false positive calls or pathogenic CNVs were missed by CMA).

Although only two cases of mosaicism were included in this study, each with approximately 50% of mosaic cells, it has been extensively reported that LP-WGS can identify low-level mosaicism with high-accuracy in different sample types, including plasma cell-free DNA for non-invasive prenatal testing (NIPT) ^8, 10, 34^ and liquid biopsy^14, 35^. In both diagnostic tests, the NGS-based approach detect mosaicism as low as 4%, mostly for aneuploidy and large copy number alterations (>5 Mb). Of note, the smallest alteration assessed in our analysis was 75 kb. However, LP-WGS resolution in clinical cytogenetics varies between studies; the sequencing method and parameters differ (i.e., single or paired-end, mean read count, and the read-length median), and there are no international guidelines for utilizing that sequencing approach in the application of CNV detection for both wet-lab and dry-lab procedures. Despite the development of new algorithms optimized the analysis of LP-WGS data, allowing an increase in resolution to be 50 kb for all types of CNVs, the most frequently reported size limit for CNV detection was 100 kb ^23^. In general, longer read lengths and paired-end sequencing provides more reliable information about the coordinates of the CNV boundaries, thus improving variant calling. That was the reason to choose this parameter and be more conservative in our analysis, but several medical centers utilize single-end sequencing most likely due to the increase in costs and sequencing time required by paired-end.

Because LP-WGS is becoming a common diagnostic approach, one aspect that stands out is its highest diagnostic yield versus CMA reported on large cohorts ^17, 18, 20, 21, 31, 33^. Nonetheless, variants of unknown significance (VUS) represent the main reason of the increased rate of additional yield, which certainly increase the challenge in CNV clinical interpretation. This particularly raise concerns for genetic counseling in both prenatal and postnatal diagnosis. Besides reviewing the prevalence of each variant in public databases (DECIPHER, DGV, ClinVar, ClinGen), a comprehensive in-house dataset including data generated from microarray and sequencing-based methods from the same population to study the pathogenicity of VUS is very useful in the clinical setting. Taking into account only aneuploidies and pathogenic CNVs, the diagnostic yield of LP-WGS (18%) was comparable to CMA (15-20%), depending on the microarray platform used ^17, 18, 20, 31^. But a limitation of the method is its inability to detect triploidy, which is particularly relevant in the context of prenatal diagnosis. Further, even though we did not observe a difference on the detection rate of LP-WGS on fetal demise samples, most likely because of our limited number, Dong et al. highlighted the requirement of a high-quality DNA for an efficient performance of LP-WGS; 6.4%, 21/328 samples from spontaneous abortions and stillbirths failed in their study ^31^.

Considering the sequencing capability of multiplexing LP-WGS, the Illumina NovaSeq 6000 platform, for example, can run up to 64, 150, 384 and 768 samples, on average of 1x coverage, on SP, S1, S2 and S4 flow cells, respectively. This demonstrates the benefit of running a great number of samples in a single sequencing slide, which ultimately lead to reduce the sequencing cost per patient. Also, a cost comparison between LP-WGS and microarray shows approximately 50% reduction in expense, being more cost-effective than any CMA platform being used in the clinical setting. It is relevant to mention though that other advantages of the sequencing approach include the requirement of low input of DNA (25 ng) when compared to CMA (200-600 ng), a significant reduction of technical repeat rates (4.6% CMA versus 0.5% LP-WGS)^23^, and it takes only one day to run an experiment compared to 3,5 days for CMA. Thus, LP-WGS decreases the experimental and labor costs in diagnostic laboratories. This is particularly relevant in the context of prenatal diagnosis since many times there is a limited amount of DNA extracted from amniotic fluid cells, which is the major cause of failure or impediment for CMA experiments.

In summary, our study demonstrates the potential use of LP-WGS to detect chromosome imbalances in clinical cytogenetics. Because of its lower cost, higher resolution, and sensitivity, the NGS-based method is a good alternative and can eventually replace CMA depending on the clinical scenario. Furthermore, our data revealed that chromosomal diseases and microdeletion/microduplication syndromes can be effectively diagnosed by LP-WGS in both prenatal and postnatal samples. The use of a commercial software facilitated testing and implementing the LP-WGS in a diagnostic laboratory.

## Data Availability

All data generated or analyzed during this study are included in this published article.

## AUTHOR CONTRIBUTIONS

Conceptualization, D.V.; A.C.V.K; C.R; Methodology, S.S.C; F.M.; P.M.P.; A.B., L.G.D.A.; M.G.P.; R.G-S; Validation, S.S.C; F.M.; P.M.P.; A.B., L.G.D.A.; M.G.P.; R.G-S; Formal analysis, P.C.M.; Data curation, P.C.M; A.C.V.K; C.R; Writing - Review & Editing, P.C.M.; D.V.; C.R.; Supervision, C.R.; Project administration, C.R.; F.M.; M.P.M.; Funding acquisition, C.R.; S-N.C.

## DATA AVAILABILITY

All data generated or analyzed during this study are included in this published article.

## ETHICS APPROVAL

This study was approved by the Ethics Committee from Institute of Bioscience - University of São Paulo (CAAE: 53093821.3.0000.5464), and an informed consent was obtained from the patients or patient’s parents for genetic testing.

